# The Virtual Parkinsonian Patient

**DOI:** 10.1101/2024.07.08.24309856

**Authors:** Marianna Angiolelli, Damien Depannemaecker, Hasnae Agouram, Jean Ŕegis, Romain Carron, Marmaduke Woodman, Letizia Chiodo, Paul Triebkorn, Abolfazl Ziaeemehr, Meysam Hashemi, Alexandre Eusebio, Viktor Jirsa, Pierpaolo Sorrentino

**Affiliations:** Aix-Marseille Univ, INSERM, INS, Institut de Neurosciences des Systèmes, Marseille, France; Department of Engineering, Università Campus Bio-Medico di Roma, Rome, Italy; Aix Marseille Univ, CNRS, INT, Institut de Neurosciences de la Timone, Marseille, France; Aix Marseille Univ, UMR INSERM 1106, Dept of Functional Neurosurgery, Marseille France; Medico-surgical Unit Epileptology, Functional and Stereotactic Neurosurgery, Timone University Hospital, Marseille, France; APHM, Hôpitaux Universitaires de Marseille, Hôpital de la Timone Department of Neurology and Movement disorders, France; Department of Biomedical Sciences, University of Sassari, Sassari, Italy

**Keywords:** neuromodulation, brain dynamics, Parkinson’s disease, Bayesian inference, model inver-sion, basal ganglia network

## Abstract

This study investigates the influence of the pharmacological nigrostriatal dopaminergic stimulation on the entire brain by analyzing EEG and deep electrodes, placed near the subthalamic nuclei, from 10 Parkinsonian patients, before (OFF) and after (ON) L-Dopa administration. We characterize large-scale brain dynamics as the spatio-temporal spreading of aperiodic bursts. We then simulate the effects of L-Dopa utilizing a novel neural-mass model that includes the local dopamine concentration. Whole-brain dynamics are simulated for different dopaminergic tones, generating predictions for the expected dynamics, to be compared with empirical EEG and deep electrode data. To this end, we invert the model and infer the most likely dopaminergic tone from empirical data, correctly identifying a higher Dopaminergic tone in the ON-state, and a lower dopaminergic tone in the OFF-state, for each patient. In conclusion, we successfully infer the dopaminergic tone by integrating anatomical and functional knowledge into physiological predictions, using solid ground truth to validate our findings.

## Introduction

Parkinson’s disease (PD) is the second-most common neurodegenerative disorder, imposing a significant socio-economic burden [1]. The pathophysiology is characterized by the degeneration of dopaminergic neurons in the substantia nigra, and the consequent depletion of dopamine in the nigrostriatal pathways [2]. Such alterations disrupt normal activity patterns, affecting brain dynamics on a large scale [3]. Accordingly, changes in the cortical dynamics are more predictive of clinical symptoms as compared to basal ganglia dynamics [4]. In turn, the symptoms are not restricted to motor abilities but, rather, involve multiple domains [5]. In line with this, structural Magnetic Resonance Imaging (MRI) has shown that the areas impacted by PD are more extensive than once believed [6]. Functional MRI (fMRI) has revealed dysfunctions in the cortico-striatal networks, with these disruptions extending to various other brain regions [7]. A magnetoencephalography (MEG) study demonstrated global changes in fast brain dynamics in PD, showing stereotyped brain dynamics as compared to controls, with the flexibility of the dynamics shrinking proportionally to clinical impairment [8]. How does the degeneration primarily occurring in the nigro-striatal pathways impact the whole brain? More specifically, how does the inability to sustain appropriate dopaminergic tone in the nigro-striatal pathways affect activities elsewhere? We aim to elucidate how effective dopaminergic tone influences large-scale brain dynamics, drawing from empirical data [9] and utilizing whole-brain models [10]. To accomplish this, we leverage data acquired from Parkinsonian patients both before (OFF state) and after (ON state) the administration of treatment with L-Dopa. Each patient has six EEG electrodes placed above the motor areas and two deep leads (four contacts each) near the left and right subthalamic nuclei (STN) [9]. We use the recently described avalanche transition matrix (ATM) to capture the brain dynamics [11]. Focusing on aperiodi bursts of activities (i.e. neuronal avalanches), the ATMs capture the probability of large-scale activities consecutively propagating across any two regions. The transition probabilities are altered in neurode-generative diseases [12, 13], and by the presence of a task [14]. This way, we have quantified the spread of the activities recorded in the electrodes implanted in patients and the EEG both in the ON and the OFF states.

In parallel, we deploy a model, known as the Dody (Dopamine Dynamics) model, that is derived from the adaptive quadratic integrate-and-fire (aQIF) model of individual neurons [10]. While originally designed for a single population of neurons, we couple the model’s equations according to three different connectivity matrices, representing excitatory, inhibitory, and dopaminergic connections, respectively. We wish to study a variable of interest that represents the dopaminergic tone, and its effect on the whole-brain dynamics. This variable represents the ability of the nigro-striatal pathways to effectively project their activities to the rest of the brain (that is, to effectively respond to a dopamine load). The evolution of local dopamine is captured by a dedicated differential equation, with one term corresponding to the increase of concentration due to dopaminergic projection and a second term capturing the reuptake mechanisms through Michaelis-Menten kinetics [15, 16], which describes the dynamics of dopamine concentration. In turn, the dopamine concentration impacts the model’s dynamics, as it is included in the equation of the membrane potential. This effect is mediated by the connectome (which we assume to be constant in the ON and OFF state) [17], and a free parameter, which is our parameter of interest to estimate, as it provides insights into the effectiveness of the stimulation. We have then projected out the simulated activities to virtual electrodes, thence providing a prediction that can be straightforwardly compared to the empirical data. In simpler terms, the model predicts how changes in dopaminergic tone affect large-scale brain dynamics. Note that these predictions are not informed by the empirical data but, rather represent theoretical arguments.

Then, we set out to put our prediction to test by inverting our model. That is, starting from the empirical data, is it possible to unambiguously infer the correct levels of Dopamine? To quantify the uncertainty of the inference, we use a Bayesian framework implemented with advanced probabilistic machine learning techniques, called simulation-based inference (SBI [18]) to efficiently invert the model [19]. In other words, we infer the posterior distribution of dopaminergic tone given the features from the empirical data. If our prediction is correct (that is, the model is simulating appropriate dynamics which is changing as a function of the levels of Dopa), we expect that the model inversion would infer higher dopaminergic tone given the data acquired in the ON state, and lower dopaminergic tone when starting from data acquired during the OFF state. We test this in each participant. The overall pipeline is shown in Fig.1.

**Figure 1:**
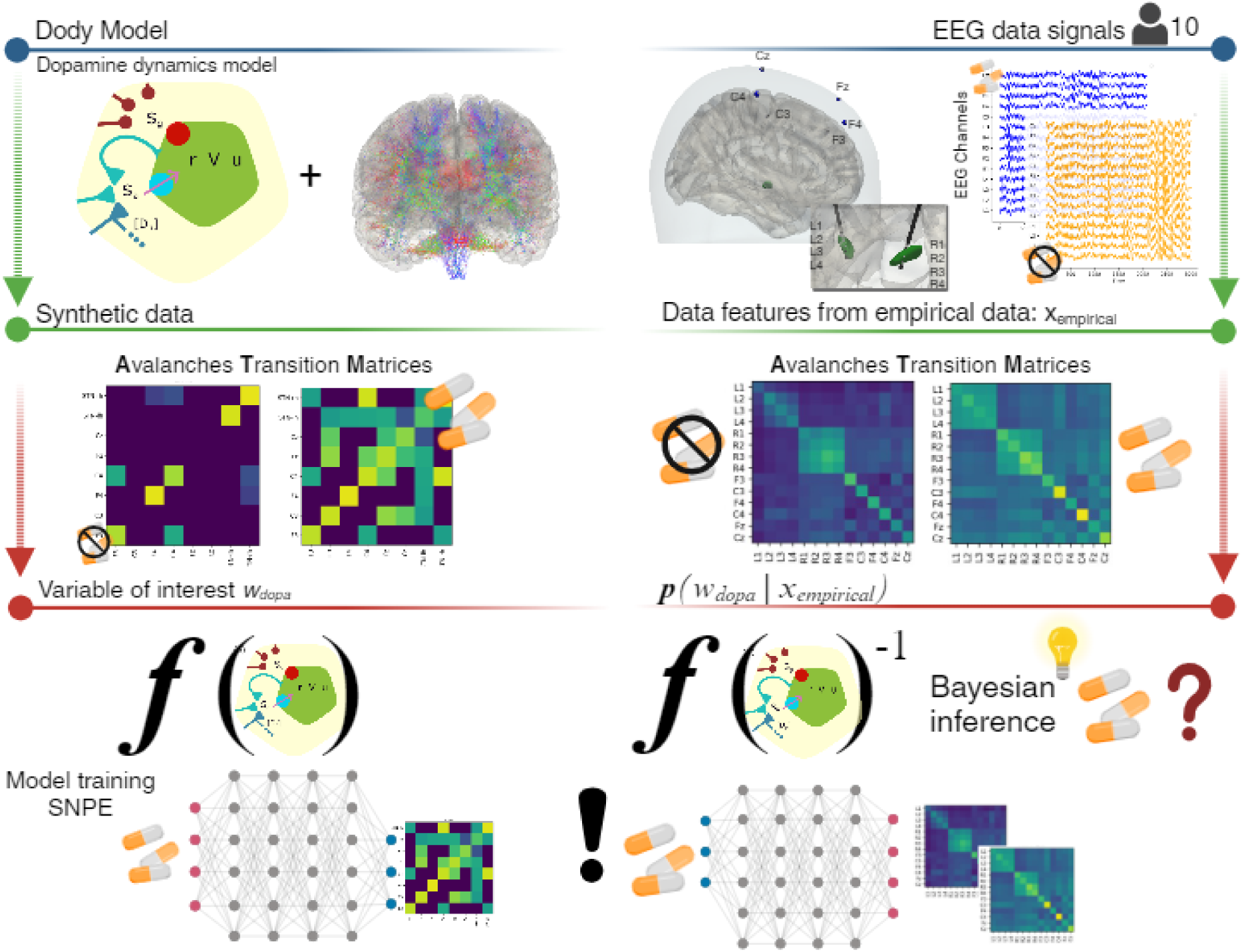
Overall Pipeline Summary. Parkinson’s disease (PD) is characterized by a progressive decline in dopaminergic neurons and a reduction in dopamine neurotransmitter levels. Progressive degeneration of dopaminergic neurons reduces dopamine content in the SN and striatum leading to PD symptoms such as tremors, instability, slow movement, and stiffness. A common treatment involves administering L-Dopa, a drug that synthesizes dopamine, though dosing can be challenging. This study aims to explore how the nigrostriatal pathways’ ability to maintain dopaminergic tone influences overall brain function. We analyzed data from EEG and deep electrodes in 10 patients with Parkinson’s, recorded before and after L-Dopa administration. To model the effects of L-Dopa in silico, we developed the Dody Model, a mechanistic neural-mass model that incorporates dopamine concentration, and Bayesian inference to infer the posterior probability distribution of dopamine concentration *w̃*_dopa_ given observed in the recordings. To do this, we used probabilistic machine-learning techniques (SNPE) to efficiently estimate an invertible function between parameters and low-dimensional data features. This way, we obtained a numerical prediction of the expected dynamics (in terms of properties of the ATMs), as observed in the deep electrodes and the EEG, given different dopaminergic tones.

## Results

### *Dody model* : generation of dopamine-driven dynamics

Starting from the model described in this work [10], where dopamine has been included, we modify the connection terms, expanding from a single-node framework to whole-brain according to three distinct layers that simulate different types of neuronal connections: excitatory, inhibitory, and dopaminergic. Each structural connection can be part of one layer (that is, it is either excitatory, inhibitory, or dopaminergic). A connection scaling factor, either *w*_dopamine_(*w_dopa_*), *w*_inhibitor_(*w_inh_*), or *w*_excitatory_(*w_exc_*), modulates each layer. In other words, if a connection is excitatory, it will be scaled by *w*_exc_, inhibitory by *w*_inh_, and dopaminergic by *w*_dopa_. The connections are shown in panel A of Fig.2. The types of connections are not merged because they play different roles in the brain. This is encapsulated mathematically as each type of connection enters the equations differently, simulating different action mechanisms.

**Figure 2:**
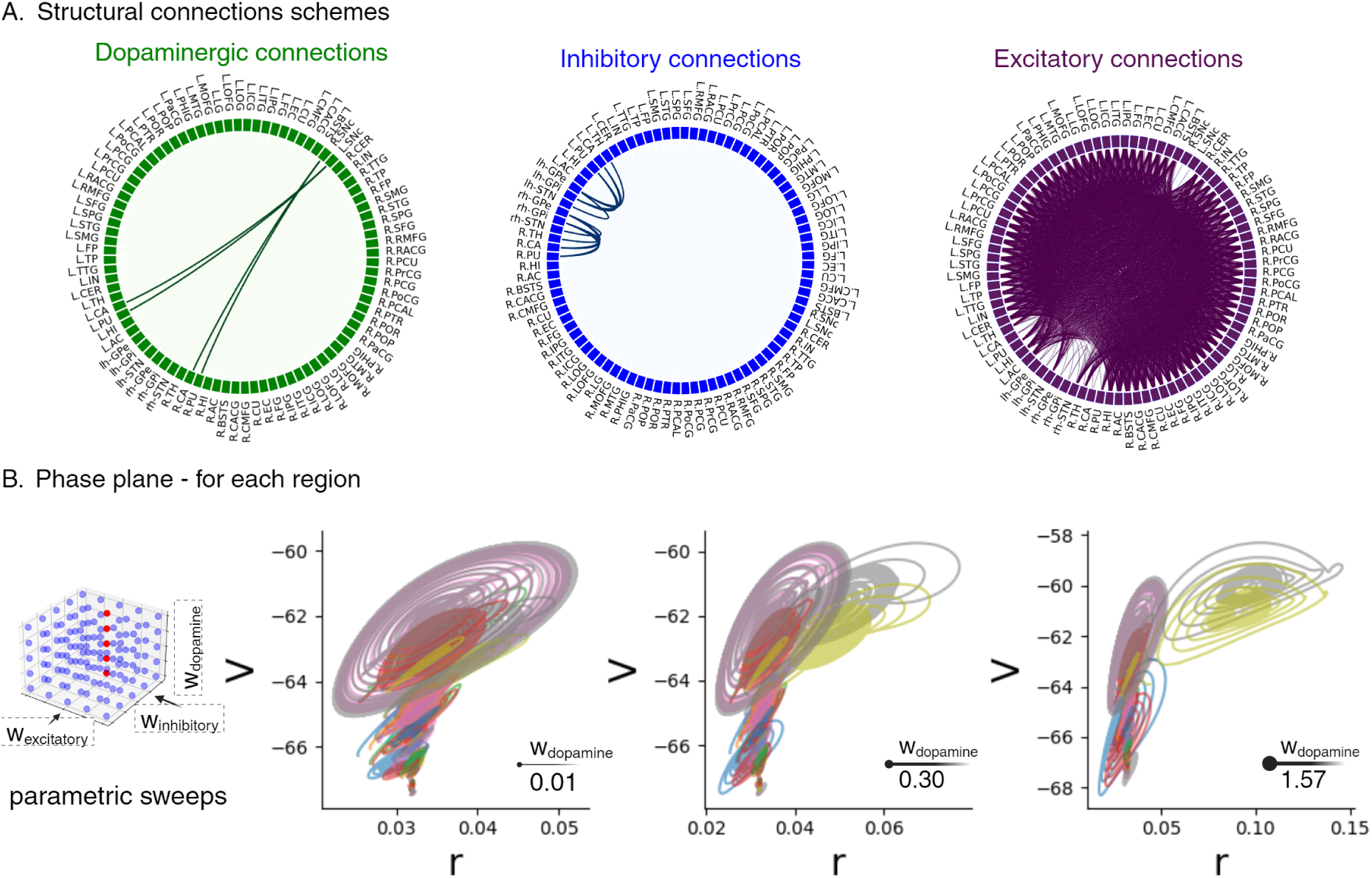
Dody simulation of the whole brain dynamics. Panel A: chord diagrams representing the three types of structural connections between 88 brain regions. Connections within these layers are scaled by three factors — *w*_dopamine_*, w*_inhibitory_ and, *w*_excitatory_. By setting precise combinations of *w*_inhibitory_and *w*_excitatory_ values, the activity of the entire network changes as shown in phase planes of the fast subsystem composed of the r-V variables (each curve refers to the dynamics of one region), namely firing rate and voltage, as a function of increasing *w*_dopamine_ (left to right).

A parameter exploration is carried out to observe the qualitative changes in the system dynamics. To understand the dynamics of the model, we use previous results at the single node level [10]. Then, we focus on the network level to understand how the interplay between the three layers of connectivity (excitatory, inhibitory, and dopaminergic) would affect the overall dynamics. For these reasons, we systematically vary the parameters (*w*_dopa_, *w*_inh_, *w*_exc_) to shift the system into a different regime. Thus, we set these values within the range that would let the whole-brain model exhibit a rich dynamical repertoire, as observed in brain recordings. In other words, we aim to simulate emerging dynamics at the whole-brain level.

Fig.2 provides an overview of the dynamics generated by the *Dody model* as a function of *w*_dopa_. This simulation aims at mimicking scenarios when patients are being administered L-Dopa.

In the next step, we project the simulated activities to the sensor space using a lead field matrix (Fig.3, panel A). This is a necessary step, going from the simulated source activities to time series that can be directly compared to the empirical data. Specifically, we project out the source activities to the EEG channels located above the motor-sensory areas bilaterally, that is F3, C3, F4, C4, Fz, and Cz. In short, the sensor-level signals are understood as a weighted linear superposition of all the source-level activities. As per the subthalamic nuclei (STN), the simulated voltages are compared directly to the lead field potential recorded by deep electrodes placed in the STNs bilaterally. Fig.3, panel B, to the left, reports the time series of the simulated voltages in OFF above and in below. Going from the bottom to the top, the first six time series correspond to the simulated EEG signals (i.e., after the projection via the lead field), and the remaining two time series correspond to the activities of the subthalamic nuclei (see methods for details).

**Figure 3:**
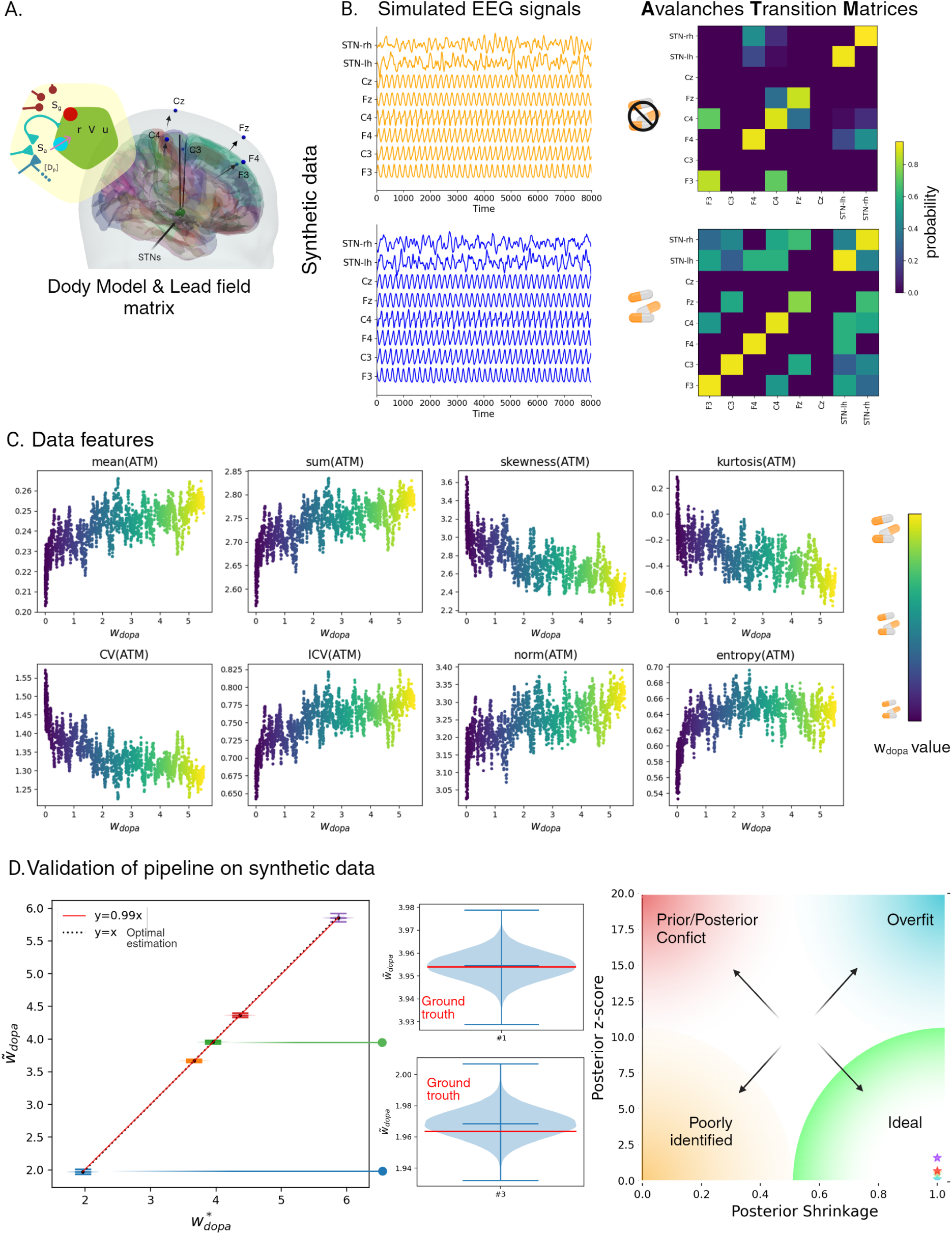
Analysis of simulated EEG signals and low-dimensional features extraction. Panel A: the simulated data are reconstructed at the sensor level for the channels in the motor frontal area, using the lead-field matrix. Panel B: the process of feature extraction from the z-score normalized EEG synthetic data, with *θ* = 1.5 as the threshold. Panel C: variations of the simulated features as a function of varying *w*_dopa_. The range of colours shows the increase in *w*_dopa_. Panel D: validation of SBI pipeline on 5 random synthetic datasets uniformly selected from an interval ranging from 0.05 to 5. The left plot illustrates the estimated values in a black dashed line, represented as *w̃*_dopa_, which are interpolated by a linear regression line that closely follows the red dashed line defined by *y* = *x*, indicating a perfect fit. The right plot displays the distribution of the posterior z-score versus posterior shrinkage, indicating an ideal Bayesian estimation (z-scores close to zero and shrinkages close to one).

To characterize the spatio-temporal dynamics over a large scale, we utilize avalanche transition matrices (ATMs), which track the spreading of nonlinear bursts. First, the z-scored signals have been binarized, and set to 1 for *z >* 1.5, and to 0 otherwise. This way, the bursts of activations are identified. Then, for each burst, the probability of two channels being consecutively recruited is estimated, obtaining an *N × N* matrix, with *N* the number of channels. These matrices are then averaged, element-wise, over the number of bursts, yielding one ATM per dopaminergic tone (i.e., the values of *w*_dopa_). Fig.3, panel B, to the right, shows the ATMs for low (top) and high (bottom) dopaminergic tone.

To extensively characterize the whole-brain dynamics, we extract features from the ATMs. We compute from the ATMs the arithmetic mean, the sum, the skewness, and kurtosis. Furthermore, we also compute from the ATMs the Coefficient of Variation, defined as the ratio of the standard deviation to the mean, and its inverse. Finally, the Frobenius norm and the entropy (*H*) of the matrix, defined as *H* = − Σ *p* log_2_ *p*, where log_2_ *p* is the base-2 logarithm of the probability distribution of signals, are also computed.

To learn a function to map from the dopaminergic tone to the expected dynamical features, we carry out multiple random brain simulations, each with a *w*_dopa_ drawn from a uniform distribution (within the ranges of the parameter exploration). Fig.3, panel C, shows the dynamical features as a function of the dopaminergic tone (*w*_dopa_). In each plot, each dot refers to the simulated feature at the corresponding value of *w*_dopa_. The scatter plots show a distinct relationship between the dopaminergic tone and the generated large-scale dynamics. Then, we perform the Bayesian model inversion starting from the empirical data (*_expt_*), recorded in each patient’s ON and OFF state. That is, based on the relationships that we found by simulating data features as a function of *w*_dopa_, we estimate the most likely *w̃*_dopa_ given a set of observed data features.

To estimate the posterior distributions of *w̃*_dopa_, we apply simulation-based inference (SBI) for efficient Bayesian model inversion [20,21]. We utilize a class of machine learning generative models for probability density estimation called Masked Autoregressive Flow (MAF) [22], trained with 3000 simulations, each with a value of *w*_dopa_ drawn from a uniform distribution [23]. The values of *w*_dopa_ and the features of the resulting dynamics are fed to the MAF, which learns an invertible function that maps the dopaminergic tone to the expected data features [18]. Then, the inversion process yields the posterior distribution of the most likely *w̃*_dopa_ values (i.e., the conditional probability of *w̃*_dopa_ given the data features).

Firstly, we test for the accuracy and reliability of the inversion using synthetic data. To this end, after training the MAF, we perform one more simulation starting from a value of *w*_dopa_ sampled from the priors. The resulting data features are given to the already-trained MAF to infer the *w̃*_dopa_. Finally, we check that the shrinkage of the posteriors around *w̃*_dopa_ correctly matches the *w*_dopa_ (i.e., the ground-truth). As shown in Fig.3, panel D, to the left, the inferred *w̃*_dopa_ (5 random values are shown as examples) converges to the ground truth. The plot illustrates that the estimated values, *w̃*_dopa_, are interpolated by a linear regression line, which closely follows the dashed black line that represents a perfect fit, indicating that the inferred parameters correspond exactly to the ground truth values. To the right, there is a zoom on two of these values. Fig.3, panel D, to the right, displays the distribution of posterior z-scores and posterior shrinkage within the Bayesian distribution (see section Methods), highlighting typical pathologies of Bayesian inference. The estimated posterior distributions for different configurations exhibit high shrinkage and low z-scores, which qualifies them as ideal Bayesian estimations. We then characterize the empirically recorded dynamics in the ON and OFF states, and then infer the dopaminergic tone from the empirical data.

### Empirical differences between patients with and without L-Dopa treatment

While analyzing empirical data, the primary objective is to differentiate the dynamics recorded during the ON state from the one recorded during the OFF state [9]. We compute, in each patient, the avalanche transition matrices from the EEG and LFP recordings acquired either during the ON condition or the OFF condition. We follow the procedure mentioned above, where each signal (either EEG or LFP) is z-scored and thresholded, and then the ATMs are computed. In summary, the time series and the ATMs, derived from the ON and OFF states, are shown for one subject in Fig.4, panel A, as an example. One can see that, in the ON-condition, the ATM contains higher transition probabilities. However, the transition probabilities are globally higher in the ON state, as shown in Fig.4 panel A, top left, for one subject. The other plots in panel B show the difference between the ON and the OFF states in every subject for all the features.

**Figure 4:**
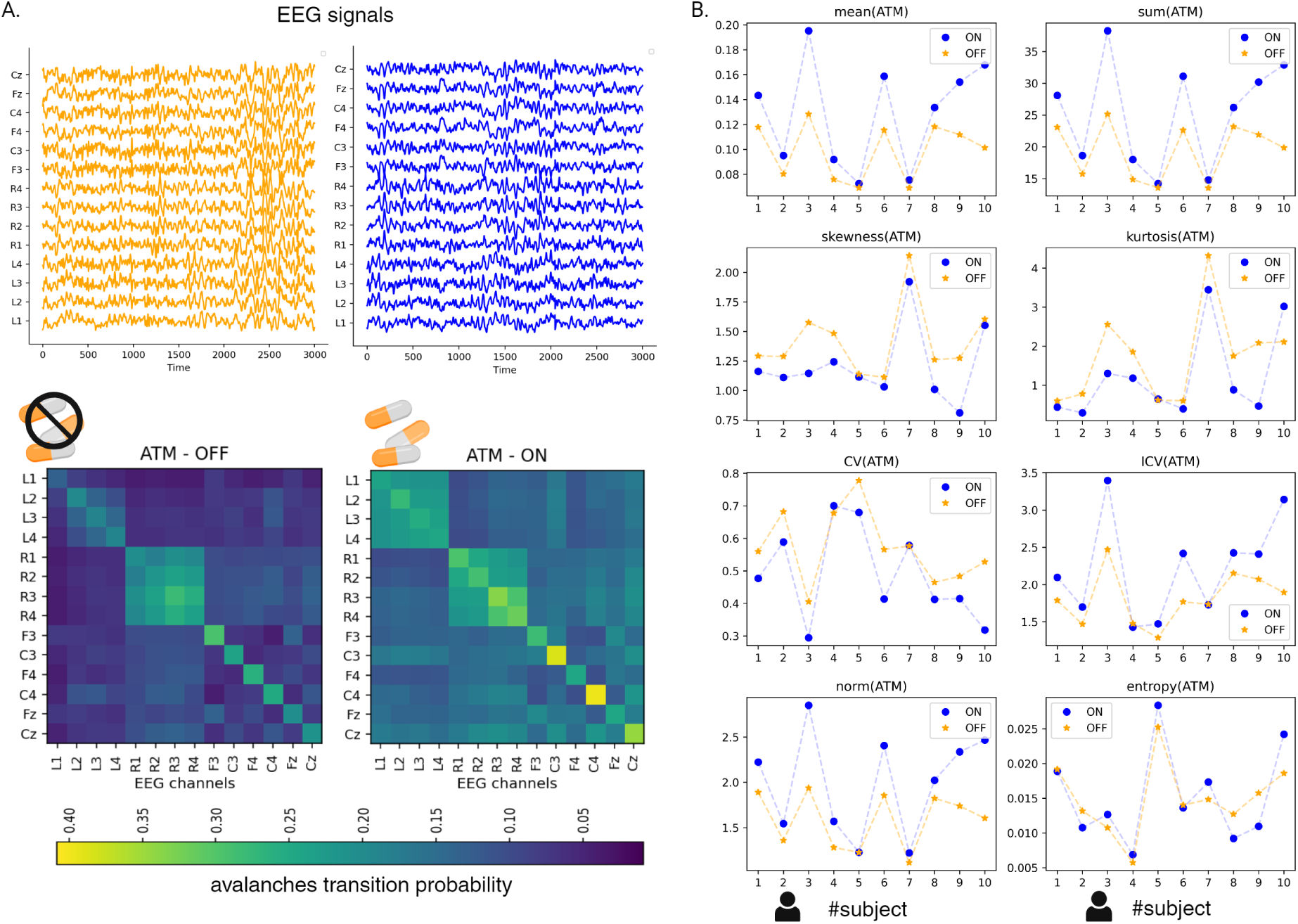
Data features from empirical data. Panel A: starting from the EEG signals, either acquired during the ON and the OFF state, the avalanche transition matrices (ATM) are computed. Panel B: a subset of the features extracted from the EEG signals and ATM matrices, for each subject in the ON (blue) and OFF (orange) states. Note that the parameters show consistent trends (with respect to the ON and the OFF states) in all the subjects.

### Model inversion and comparison of the synthetic and the empirical dynamics

Finally, the empirical data features (during the ON or the OFF state) are fed to the trained MAF that yields the posterior distribution for the ON and the OFF states. In other words, for each patient, we obtain the posterior distribution for *w̃*_dopa_ given the data recorded during the OFF state (orange) and the ON state (blue), as shown in Fig.5 panel A. For all patients, the inversion infers a lower dopaminergic tone when starting from data acquired during the OFF state and, conversely, a higher dopaminergic tone when starting from data from the ON state. We regard the ON and OFF states as ground truths, since L-Dopa had been orally given to all the participants before the ON-state recording, likely changing the effectiveness of the dopaminergic stimulation. We use the Wasserstein distance to quantify the differences between the distributions of the estimated *w̃*_dopa_ in the two conditions (Fig. 5, panel B). The Wasserstein distance between the posteriors inferred from the ON and the OFF states is always greater than 0, implying that the two distributions are significantly different. Furthermore, we report the shrinkage of the posterior distributions as compared to the priors (that is, the reduction of uncertainty about the “true” *w*_dopa_ provided by the model inversion, or the level of information in the posteriors as updated from the priors). The shrinkage for all distributions is close to 1, indicating that there is a significant reduction in the uncertainty of dopaminergic tone. Furthermore, we consider the clinical improvement observed in patients in the ON and the OFF conditions, as measured by the Unified Parkinson’s Disease Rating Scale (UPDRS), a comprehensive clinical tool used to quantify the severity of PD symptoms. Specifically, the UPDRS Part III (UPDRS-III) focuses on the motor examination and is pivotal in assessing the motor function of PD patients. For each patient, the differences of the UPDRS-III in the ON and OFF conditions are computed and correlated to the distance between the simulated ATM matrices (generated starting from the inferred *w̃*_dopa_ in the ON and the OFF states (*r* = 0, 49, p-value= 0.09), as shown in panel C of Fig. 5. However, the preliminary data suggest that the generated feature might hold behavioral significance. As a second step, we aim to directly compare the dynamics generated by the numerical simulations in the ON and the OFF states with the corresponding observed empirical dynamics. This step is taken to explore the realism of the generated dynamics beyond the ability to correctly support the model inversion. As shown in Panel D of Fig. 5, the analyses focus on the relationship between the power spectra of the simulated and empirical time series derived from the cortical channels (left) and the STNs (right). Firstly, we compute the Spearman correlation between the simulated differences (that is, the frequency-wise differences of the power spectra in the simulated ON -OFF states) and the corresponding empirical ones. The synthetic data are generated by re-simulating the model using the most likely *w̃*_dopa_ in the ON and the OFF states, respectively, averaging over 10 simulations, where the time series are different across simulations due to the dynamical noise. The first row of panel D shows, to the left, the Spearman correlation, averaged across cortical channels, between the ON-OFF differences in the synthetic data and the ON-OFF differences in the empirical data for each subject. A similar scheme is used to the right to report the results for the STN (see methods). These results point out the capability of the model to generate realistic variations, effectively mirroring the empirical data. Notably, changes in the dynamics of the STN in the model arise non-trivially through network effects, since there are no direct dopaminergic connections to the STN.

**Figure 5:**
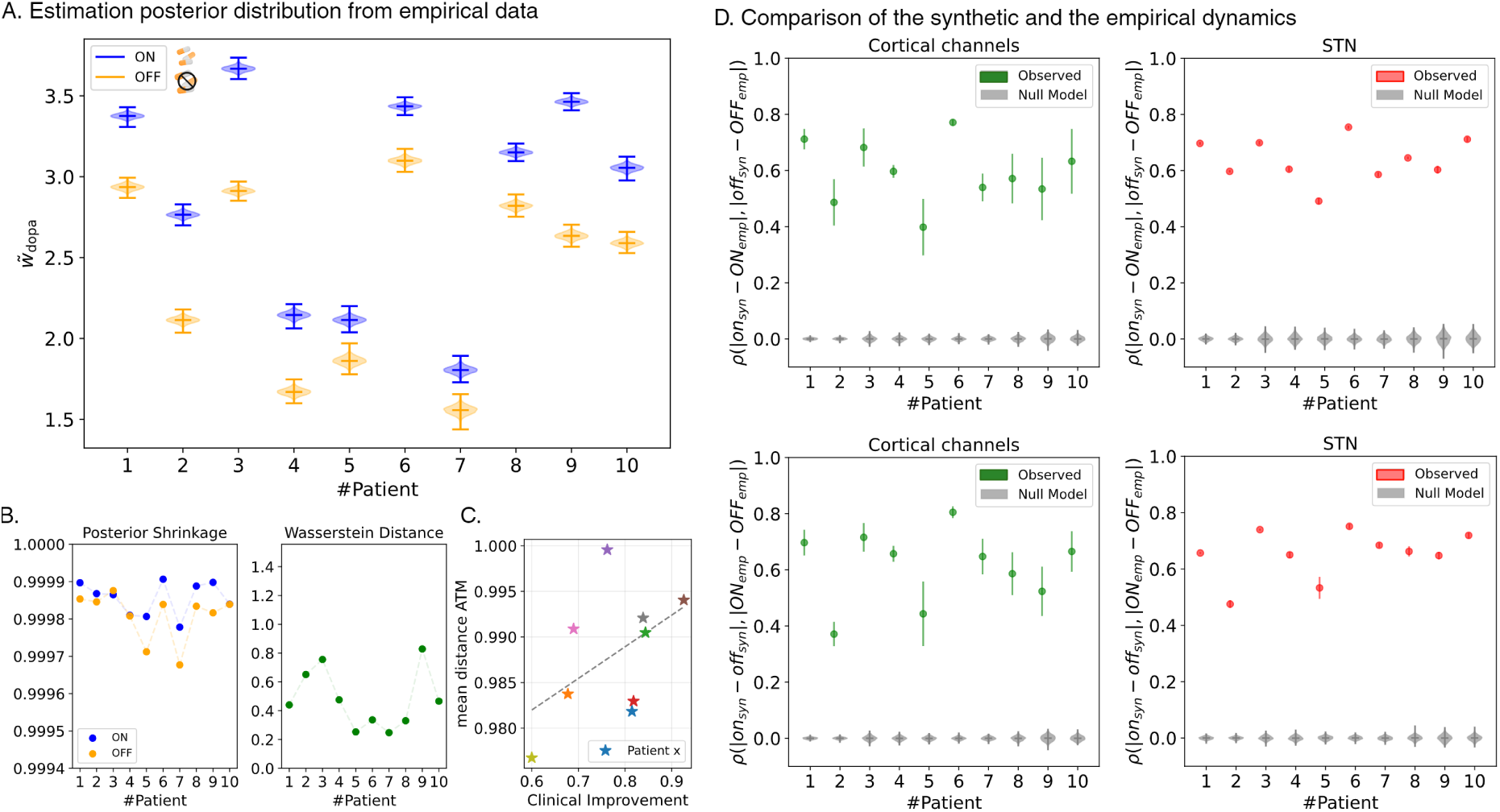
Estimation posterior distribution from empirical data. Panel A: the estimated posterior distributions, for each patient. Higher values are observed in the ON state compared to the OFF state for all subjects. Panel B: the posterior shrinkage and the Wasserstein distance to compute the uncertainty of the dopaminergic tone and the distance between the two distributions, respectively, indicating the reliability of the Bayesian estimations. Panel C: Scatterplot showing the clinical improvement (UPDRS-III) between the ON and OFF conditions for each patient (each point represents a patient) and to the distance between the simulated ATM matrices (generated starting from the inferred *w̃*_dopa_ in the ON and the OFF states (*r* = 0, 49, p-value= 0.09). Panel D: Comparison of the synthetic and empirical dynamics across Cortical channels (left panels) and Subthalamic Nuclei (STN) (right panels). The first row displays the Spearman correlation *ρ* between the differences in power spectra of the ON and OFF states, as computed for the synthetic and empirical data. For all subjects, the observed correlations (green for cortical channels, red for STN) are significantly higher than those expected from the null model (gray markers), indicating that the model reliably captures the ON-OFF variations observed in the empirical data. The second row further validates the model by comparing the ON-state differences (synthetic vs. empirical) to the OFF-state differences (synthetic vs. empirical). Consistent correlations across subjects demonstrate that variations in the ON state are mirrored in the OFF state, supporting the model’s robustness in reproducing state-specific dynamical features. In all cases, p-values*<* 0.001.

Analogously, the second row of panel D shows the Spearman correlation between ON state differences (synthetic vs. empirical) and OFF state differences (synthetic vs. empirical). The results reveal a positive correlation and a consistent trend across patients, meaning that small differences between the ON states correspond to small differences between the OFF states, and large differences in the ON states are similarly mirrored in the OFF states. This confirms that the model captures empirical ON-OFF variations at the cortical level (to the left) and for the STNs (to the right). To validate all the correlations reported above, we randomize the power spectra by shuffling the frequency bins, to estimate a null distribution of the correlations. This systematic comparison highlights the model’s ability to reproduce realistic and consistent changes in the dynamical features of both cortical and STN signals, well above the chance level.

As a last analysis, we explore the time series of the dopamine concentrations [*Dp*]*_e_* (see equation 6) for different *w̃*_dopa_ values, as shown in figure S2 of Supplementary Materials. Striatal regions show higher levels of Dopamine in the ON state compared to the OFF state.

## Discussion

In this article, we set out to implement a model to infer dopaminergic tone in individual patients from a combination of EEG electrodes and intracranial deep leads before and after L-Dopa administration. Simulating data using the model and then inverting it using deep neural networks, we successfully infer the dopaminergic tone starting from empirical data acquired before and after the intake of L-Dopa.

The Bayesian framework allows us to integrate the background knowledge as a prior distribution, and reveal the mechanism(s) starting from macroscopic measurements. Identifying the patient’s state, either ON or OFF, is not based directly on learning statistical features but, rather, on the inference of a biologically plausible mechanism [24]. This is opposed to other approaches that focus on predicting data patterns based on empirical observations, without providing insights into the causal mechanisms.

The *Dody model* is a neural mass model based on the adaptive quadratic integrate-and-fire model (aQIF), with some differences from the previous models used thus far [10]. The neural mass has been modified from [25] to include an additional variable capturing the local dopamine concentration. Hence, each connection affects the neural masses differently, according to its nature, that is, dopaminergic, excitatory, or inhibitory. In other words, the neural masses are coupled according to the empirical connectome, where connections can be excitatory, inhibitory, or dopaminergic. This model encapsulates a wealth of well-established information about the specific role of each connection (i.e., the directionality and its type), which has been typically disregarded in large-scale models thus far [26, 27]. To do so, each tract, as measured from the tractography, is considered with directionality and with a specific functional role (as an example, the tract from *i* to *j* might be inhibitory while the tract *j* to *i* excitatory). The individual structural connectomes are not available. Therefore, unlike typical virtualization pipelines, we build one generic model to perform inference on each patient. As explained, the coupling is done utilizing a connectome computed as the edge-wise average of 10 healthy subjects extracted from the Human Connectome project [28]. This particular design speaks to the validity and the generalizability of our pipeline. In fact, in this work, we aim to capture the functional deficit of the nigro-striatal pathways which is the primum movens in each Parkinsonian patient [29]. Accordingly, our model simulates the expected changes in terms of large-scale brain dynamics as a function of the nigro-striatal activities. We then set out to provide preliminary validation of the model’s predictions by inferring lower/higher levels of Dopamine from the data acquired in the OFF/ON state. To this end, the same model is inverted starting from data from each of the 10 patients, either in the ON or in the OFF states, and the inversion is successful in each, as shown by the shrinkage of the posteriors. This might be interpreted as the fact that the model, and the features used for the inversion, capture the effects that L-Dopa exerts on the dynamics of patients in general. This mechanistic explanation constitutes a hypothesis of the effect of L-Dopa on the corresponding large-scale activities. Based on this evidence, the forward solutions of our model provide a prediction that is not yet personalized but reproduces the expected changes of a generic set of features expected as a function of changing Dopaminergic tones [30]. However, the model successfully distinguishes each patient’s ON and OFF conditions. In other words, our model provides an answer to the question ”How do changes in the ability of the nigro-striatal pathways to uptake L-Dopa, process it, and release dopamine affect brain dynamics, as observed from a change in data features from large-scale data?” As explained, the inference of the levels of Dopamine should not be understood as a way to tell apart the ON and the OFF states (which is easily attainable from data features alone) but, rather, as a check of the validity of the model. A more comprehensive validation of the model would require an interventional study to verify that the relationship between Dopa and large-scale dynamics holds across the range of values explored by the model. Here, we exploit the fact that we have two different levels of Dopamine available, as they were administered to patients. Hence, the inversion is done to check the ability of the model to correctly and unambiguously distinguish the whole-brain dynamics for the two levels for which we have the ”ground truth”. However, as explained, the model’s predictions spann a much larger range of values (which we cannot test).

Concerning the data features that we utilize, we focus on the topology of the spread of large-scale aperiodic bursts [11, 31]. This approach is complementary to most traditional methods, which focus on the presence of synchronization or the dynamics of local bursts, typically in the beta band (13-30 Hz) [32–37]. In addition to calculating various features from the EEG signals, we utilize the recently described ATMs to capture the spatiotemporal spreading of each such perturbation [11]. Of note, we find that the ATMs are at once informative about the large-scale dynamics yet low-dimensional, as we can accurately estimate the posterior distribution from these features (e.g. the mean, the kurtosis, etc.), and observe the corresponding differences between the ON and the OFF states at the individual level. The experimental setting, contrasting each individual before and after the administration of L-Dopa, allows for establishing a causal relationship between the administration of L-Dopa and the changes in the dynamics over the large scale. Nevertheless, revealing a true causal mechanism requires considering inference on multiple causal factors followed by a comparison to the evidence. This aspect requires a thorough investigation and has not yet been demonstrated using SBI. Furthermore, we demonstrate the possibility to define cross-modality connectivity matrices and deploy them for inference, since some transition probabilities are computed starting from deep electrodes and going to EEG electrodes, or vice-versa.

In our scenario, we want to simulate the effects of varying levels of dopaminergic tone (represented by different values of *w*_dopa_) on the data features. This allows us to estimate the posterior distributions of *w̃*_dopa_, determining the probability of a specific dopaminergic tone given the empirical features observed.

In particular, to estimate the posterior distributions of the parameter of interest, we apply SBI for efficient Bayesian model inversion. This approach is necessary because the calculation of the likelihood function at the whole-brain scale is often intractable, and Markov chain Monte Carlo (MCMC) might be inapplicable for non-parametric sampling. Note that, in the Bayesian setup, the generative model is represented by the joint distribution of model parameters and data, presenting computational and convergence challenges arising from the high dimensionality of the data space, despite the low dimensionality of the parameter space. By harnessing the complexity of mechanistic models and using low-dimensional data features, SBI allows us to efficiently infer the underlying dopaminergic tone from the observed data, providing a robust and neurophysiologically grounded understanding of the system. By using state-of-the-art probabilistic machine-learning tools for probability density estimation, such as MAF, the SBI is an efficient approach, as it relies only on forward model simulations. Moreover, since SBI requires low-dimensional data features for training, the ATM proved useful, since they capture dopamine-induced changes (as validated by the inversion starting from empirical data). Furthermore, the changes in the ATMs are predictive of clinical impairment. However, these results are to be regarded as explorative, given the low numerosity. The applicability of SBI applied to our model is confirmed by the close alignment of predicted and empirical data features, with the model showing particular sensitivity to changes in the dopaminergic tone, capturing a general mechanism, and linking the effect of medical treatment to whole-brain dynamics, which applies to each participant. As confirmation of the reliability and effectiv-ness of the inversion, we calculate the shrinkage of the posteriors from the priors, to quantify the level of information held therein, starting from the priors. By comparing the shrinkage of the ON and OFF states, we can assess the consistency and reliability of the distributions [38].

Finally, we directly compare the differences (ON vs. OFF) in the power-spectral densities of the empirical and simulated (starting from the inferred *w̃*_dopa_ in the ON and the OFF states) timeseries.More specifically, we first compute the Spearman correlation between the frequency-wise differences of the power spectra in the simulated ON-OFF states and the corresponding empirical ones (Fig.5, panel D). Secondly, we also compute the Spearman correlation between frequency-wise differences of the power spectra (synthetic vs. empirical) in the ON state and the corresponding differences in the OFF state.

Notably, we observe a significant correlation between the differences, indicating that the simulated dynamics capture the empirical effects of Dopamine over the large-scale dynamics.

Additionally, by examining the temporal evolution of the distribution of dopamine concentration, we obtain further confirmation of the validity of our approach. This is evident as we observe distinct features when simulating the ON states as compared to the OFF state. While it cannot be checked with the data at hand, our model predicts that patients with lower levels of *w*_dopa_ will converge to lower concentrations of L-Dopa in the Striatum over time, as opposed to patients with higher levels of *w*_dopa_, which will steadily display more L-Dopa. The differences, however, are patient-specific and mediated by network effects. In other words, our model predicts the ability of the nigrostriatal pathway to affect whole-brain dynamics. These theoretical predictions, however, can only be partially tested in the empirical data, as these experiments are prohibitively difficult or impossible to perform. This is the advantage of virtual brain modeling, which allows us to test our hypotheses *in silico*. As explained, we compare the differences in the power spectra in the ON-OFF states in the simulated and empirical data in the STN (where deep electrodes were placed). We show that the correlation is higher than expected by chance, corroborating the validity of the proposed approach. Note that the effects of L-Dopa in the STN necessarily represent network effects, as no direct Dopaminergic connections are hinging on the STN. Future advancements might involve setting heterogeneous *w*_dopa_ values for each hemisphere, enhancing the representation of dopaminergic tone variability across brain hemispheres. This strategy could refine clinical interventions (given the clinical asymmetry typical of PD), facilitate comparative studies across brain hemispheres, aid in developing new biomarkers, and improve simulation models [30]. Importantly, it could enhance research into Parkinson’s Disease by allowing for personalized treatment strategies. This is because accurately estimating the dopaminergic tone based on EEG data dynamics can help predict a patient’s clinical state [9]. On the one hand, more biological details might be added to the model. On the other hand, more parameters might generate degeneracy and make causal estimation more challenging. Hence, further studies show refine this approach and find an optimal trade-off. Finally, further studies should specifically address the problem of dyskinesias or uncontrolled involuntary movements.

Our work is complementary to most of the current modeling literature in PD, which primarily focuses on the simulation of activities in the beta range [39, 40]. We focus on aperiodic activities instead, which have only received limited attention in Parkinson’s disease thus far. Furthermore, we focus on large-scale dynamics and efficient parameter estimation with the associated uncertainty, rather than directly utilizing a cost-function to fit the model [41, 42]. Other modeling works in PD, for example in the case of the Virtual Deep brain stimulation, took a multi-scale simulation approach in order to simulate the effects of deep brain stimulation [43, 44]. However, the scope of our model is different, as we provide an explicit account of the concentrations of Dopamine, which brings us closer to pathophysiological mechanisms [45]. In other words, our model explicitly encapsulates physiological knowledge about the functioning of dopamine and its effects on neural activities. Finally, our model aims at describing the whole brain, unlike previous works which represented the cortex as a single node. In conclusion, our work analyses EEG/LFP data from PD patients in the ON and the OFF states focusing on the aperiodic bursts, and leverages a newly designed mechanistic model that explicitly includes dopamine dynamics.

## Materials and Methods

### Participants, EEG Data Acquisition and Processing

We collected resting-state data from 10 patients with Parkinson’s disease, and monitored both ON and OFF levodopa medication [9]. Each session was 2 minutes long, employing deep stimulation electrodes and EEG electrodes strategically placed in the motor areas on both sides. This setup enabled simultaneous recordings from both the subthalamic nuclei (STN) and the motor cortex. Specifically, the data includes time series from 14 channels: eight local field potential (LFP) contacts (L1, L2, L3, L4, R1, R2, R3, R4) that map to the left and right STN complemented by six EEG channels (F3, Fz, F4, C3, C4, Cz). The local ethics committee approved the study, and all participants provided written informed consent.

Signal preprocessing was performed using the Fieldtrip toolbox [46]. The continuous EEG signal was first high-pass filtered at 1.3 Hz with a Hamming window, using a “two-pass” direction, and a Butterworth filter type. Subsequently, it was down-sampled to 512 Hz and epoched into 4-second epochs. The signal underwent visual inspection to remove noisy epochs [47].

### Tractography Data

Minimally preprocess diffusion and structural MRI data from 10 subjects of the human connectome project [48, 49] is used to calculate the structural connectomes. The diffusion imaging data is processed with the MRtrix3 toolbox [50]. Multi-shell multi-tissue constrained spherical deconvolution with group averaged response function [51] is used to estimate fiber orientation distributions per voxel. After intensity inhomogeneity correction [52], we use probabilistic tractography with anatomical constraint [53] to generate 5 million tracts per subject. Tracts are weighed using the SIFT2 algorithm [54]. We use the multimodal registration method as implemented in the Lead DBS toolbox [55] to transform the subcortical structures (GPe, GPi, STN and RN) of the DISTAL atlas [56] into the subject space. The subcortical regions together with the cortical Desikan parcellation [57] serve as the mask to group weighted tracts and create the structural connectome. To compute the lead field matrix we process the MNI152 template head with the Freesurfer recon-all [58] pipeline to obtain the cortical, as well as the 3 boundary element model (BEM) surfaces (inner skull, outer skull, and outer skin surface). We use the MNE toolbox [59] to fit a standard 10-20 EEG cap onto the skin surface of the MNI152 template head by manually setting the fiducial points (right and left pre-auricular point and nasion). Vertices of the cortical surface are used as neural electric dipoles. Together with the EEG locations and the BEM surfaces, we compute the electric forward problem using the OpenMEEG toolbox [59].

### *Dody model* for Whole Brain Simulation

In our study, we employ the *Dody model* introduced here [10] for the first time. Initially implemented to simulate the dynamics of a single node, it has been extended to simulate the whole brain activity. The model is based on neural masses designed from the adaptive quadratic integrate-and-fire model, thence incorporating neuromodulatory dynamics into its differential equations. To simulate dopamine dynamics, which is relevant in the context of Parkinson’s disease, equation 6, constituted of two terms, the first corresponding to the afferent dopaminergic input and the second to the reuptake mechanisms described with Michaelis-Menten formalism, is added to the system, as:

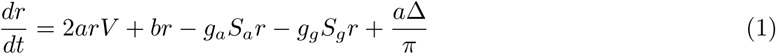

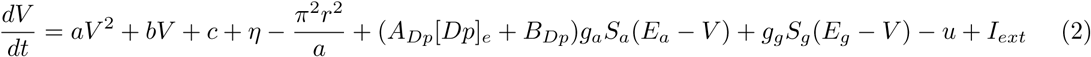

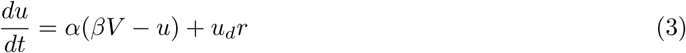

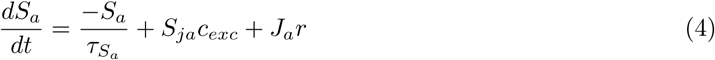

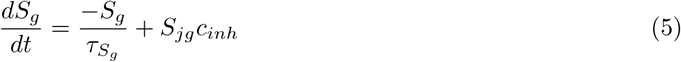

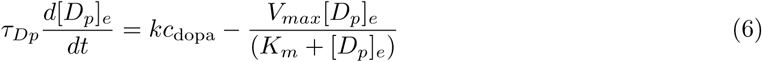

We focus our attention on the terms *c*_exc_, *c*_inh_, and *c*_dopa_, which are involved in equations (4), (5) and (6), respectively. The connection-specific terms *c*_exc_, *c*_inh_, *c*_dopa_ are informed by anatomical data from scientific literature, understanding each brain connection as either excitatory, inhibitory, or dopaminergic. The main diagonal of each of these terms represents local connectivity, while the off-diagonal elements represent long-range connectivities. The terms are modulated by a parameter *g*^∗^ through the following relations:

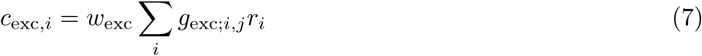

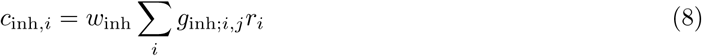

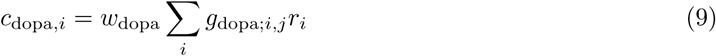

where *g*^∗^ is given by the connectome weights. In particular, cortical connections have been considered excitatory. The cortex-basal ganglia-thalamic loop has been modeled taking into account the direct, indirect, and hyperdirect pathways [60], as reported in Fig. S1 of Supplementary figures. The simulations were run for 10 seconds by numerically integrating the system of equations, through the modified Heun method for stochastic differential equations [61, 62]. The first second was discarded as a transient.

### Estimation of Avalanche Transition Matrices and Data Features

To explore the dynamics of brain activity, we analyze EEG data signals. Firstly, each signal amplitude is standardized using the z-score and then binarized, such that any time point exceeding a threshold of two standard deviations (*|z|* = 2) was marked as 1 (active), and all others as 0 (inactive) to compute avalanche transition matrices as described in this work [9, 11].

During our analysis of EEG data and the estimation of neuronal transition matrices, a key focus is on evaluating the impact of levodopa medication on the features. We calculate an avalanche-specific transition matrix (ATM), where each element (*i, j*) represents the probability that region *j* is active at time *t* +1, given that region *i* was active at time *t*. This relationship signifies the probability of sequential recruitment of any two regions by an avalanche. For each subject, we produce an average transition matrix by averaging edge-wise across all avalanches and then symmetrizing it. In our data analysis pipeline, we explore how avalanches propagate between different brain regions using ATMs. Specifically, we assess the ATMs under two different conditions for each patient: with levodopa medication (ON) and without it (OFF). This comparison sheds light on the dynamics of perturbations spreading between brain regions affected by different levels of dopaminergic stimulation. Comprehensive tests to validate the ATMs and computational details are reported in this previous work [9]. Simultaneously, avalanche transition matrices are calculated for synthetic data as well. Starting from the simulations, the firing rates of individual brain regions are translated to the positions of the six considered electrodes using the lead field matrix. Since the position of the deep electrodes is not available, they have been approximated directly as the z-scored activities, without projecting it through the lead field. At the end of the procedure, 8 signals are generated. Each signal is z-scored, and then, similarly to the procedure described above, active regions are identified to calculate the ATM as with the empirical data. However, this time we do not take the absolute value of the signals.

### Model Inversion and Dopaminergic Tone Parameter Inference

The Bayesian approach offers a principled way for making inference, prediction, and quantifying uncertainty in the decision-making process by integrating information from anatomical, clinical, and mathematical knowledge [63]. Parameter estimation within a Bayesian framework involves quantifying and propagating uncertainty through probability distributions placed on the parameters (the prior), which are updated with information provided by the data (the likelihood) to form the posterior distribution [19, 64]. However, accurate and reliable Bayesian inference from noisy brain data is challenging due to the high dimensionality of the data, the complex effects of brain networks, and the non-linearity involved in spatio-temporal brain organization. In particular, the calculation of the likelihood function is typically intractable, rendering MCMC sampling inapplicable [18]. In this case, we can treat the virtual brain models as a stochastic simulator that generates synthetic data similar to the empirical data, enabling inferences to be made without requiring access to the likelihood function [21, 23].

Using this framework known as Simulation-based Inference (SBI [21,23]), the *Dody model* is treated as a stochastic simulator, that is the generative model necessary to conduct Bayesian inference on dopaminergic tone, which is informed by anatomical data, to predict the features extracted from observed EEG data. To accomplish this, we train a class of machine-learning generative models known as Normalizing Flows (NFs [22]) to learn the relationship between low-dimensional data features and the parameters of an approximated posterior distribution. NFs are a family of generative models that convert a simple base distribution (e.g., the prior) into any complex target distribution (e.g., the posterior), where both sampling and density evaluation can be efficient and exact [65].

In more detail, our objective is to establish plausible probability distributions for the *w*_dopa_ parameter in the *Dody model*, considering that the output distributions of our model should best explain a given set of experimental data features. Considering *w*_dopa_ as the known parameter (1-dimensional) and a collection of *n* data observations, denoted by the variable *y_expt_*. The dynamical system analysis (previously conducted) provides insight into the reasonable range of parameter values, allowing us to define initial (prior) probability distributions *p*(*w*_dopa_), as a uniform distribution truncated between [0.05, 5]. Using Bayes’ rule, the posterior distribution of the parameter values given the data is defined by the following equation:

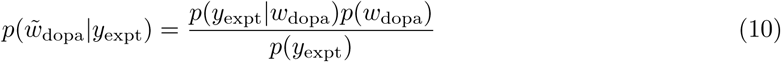

where *p*(*w̃*_dopa_) denotes the prior placed on *w*_dopa_, and the likelihood *p*(*y*_expt_*|w*_dopa_) is the probability of our model generating the data features *y*_expt_ given the parameter *w*_dopa_, and *p*(*w*_dopa_*|y*_expt_) represents the posterior distribution that we aim to estimate.

In this work, we use SNPE (Sequential Neural Posterior Estimation [23]), a tool enabling efficient and flexible simulation-based inference on complex models without requiring access to likelihoods. SNPE dynamically refines the proposals, network weights, and posterior estimates to learn how model parameters are related to the observed summary statistics of the data. We run SNPE using a single round to benefit from an amortized strategy (at the subject level), which can then be applied immediately to new data without needing to be retrained [21]. In this framework, we use Masked Autoregressive Flow (MAF [22]), which supports reversible nonlinear transformations and allows for highly expressive transformations with minimal computational cost.

To perform SNPE, three key inputs are required [21, 23]:

1. A prior distribution describing the biologically plausible values of *w*_dopa_, which modulates the dopaminergic tone influencing the ON and OFF states.
2. A mechanistic model that simulates the large-scale neural activity associated with a specific value of *w*_dopa_.
3. A set of low-dimensional yet sufficiently informative data features, focusing here on the EEG recordings.

We train the MAF using a budget of 3000 simulations with random parameters sampled from the prior. The set of data features extracted from source reconstructed EEG includes summary statistics of EEG signals and of Avalanche Transition Matrices (ATM), such as kurtosis of each signal, and the functional connectivity. From the ATMs, we compute the sum, arithmetic mean, skewness, kurtosis, the Coefficient of Variation (defined as the ratio of the standard deviation to the mean, and its inverse), the Frobenius norm, and the entropy (H) of the matrix, defined as *H* = -∑ (*p* log_2_(*p*)) where log_2_(*p*) is the base-2 logarithm of the probability distribution of signals. After training is complete, the posterior distribution for new observations or empirical data can be swiftly evaluated by performing a forward pass in the trained MAF. Notably, this process does not require the model or the data features to be differentiable. Each model simulation and posterior sampling took around 60 seconds and 30 seconds, respectively. All steps were performed on a Workstation DELL Precision 7820 Tower with 2 Intel Xeon Silver 4214R. To run SNPE, we used the public sbi toolbox [20].

Before running SNPE on the empirical data, we first validate the approach using synthetic data generated with known ground truth values of dopaminergic tone, and with subject-specific structural connectomes to ascertain the accuracy of the *w*_dopa_ estimations (see Fig. 5). The plot of z-score versus posterior shrinkage for the estimated posterior indicates an ideal Bayesian estimation for different values of *w*_dopa_.

### Data Features

To run SBI, we identify specific data features derived from synthetic EEG data, as follows: the sum and arithmetic mean of the ATM; the exponential of Skewness and Kurtosis (evaluating the asymmetry and tailedness of the ATM distribution around its mean value). The Coefficient of Variation and its inverse, which are defined as the ratio of the standard deviation to the mean, providing a normalized measure of dispersion for the ATM elements. The Frobenius norm derives a scalar magnitude of the ATM. The Shannon entropy of the ATM is computed as *H* = -∑ *p* log_2_ *p* where log_2_ *p* is the base-2 logarithm of the probability distribution of signals. For the training phase, the data features are smoothed using a moving average with a window size of 150 sec. Subsequently, a linear regression is performed to obtain the value of the feature for each *w*_dopa_.

### Evaluation of Posterior Fit

To evaluate the accuracy and reliability of the Bayesian inference using synthetic data, we compute the posterior z-score (denoted by *z*), against the posterior shrinkage (denoted by *s*):

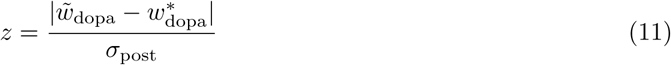

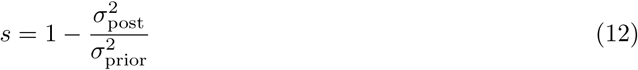

where the z-score is defined as the absolute value of the difference between the estimated and true values of the parameters of interest (*w̃*_dopa_, 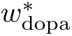 respectively) divided by the standard deviations of the posterior *σ*_post_, and the posterior shrinkage is given by 1 minus the ratio of the variance of the posterior 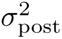 to the variance of the prior 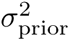.

The z-score measures how much the posterior distribution is centred around the true value, whereas the shrinkage quantifies the reduction of uncertainty over the estimate around the true value of the posterior distribution with respect to the initial prior distribution. Ideally, the distribution of posteriors derived from prior predictive observations should converge towards small z-score and large posteriors’ shrinkage for each parameter component. By plotting the posterior z-score (*y* -axis) against the posterior shrinkage (*x* -axis), a concentration towards the lower right suggests an ideal Bayesian estimation [19,66].

To study the correlation between the simulated data and the clinical improvement, we first compute the distance between the ATMs simulated in the ON and the OFF states, defined as 1 minus the absolute value of the mean of the edge-wise differences. This is correlated, across subjects, to the difference in the UPDRS scores in the ON and the OFF states. The result was validated utilizing a null distribution obtained by shuffling the averages of the differences between matrices.

As explained, we compared the ON-OFF differences in the synthetic and empirical data for each subject to compare the model’s performance to effectively capture the differences in the time series. To compare the data we proceed as follows: 1) compute the power spectra of the signals; 2) subtract (frequency-band wise) the power spectra of the signal in ON from the empirical STN in OFF, and subtract the power spectra of the simulated STN in ON from the empirical STN in OFF (i.e. with the simulations performed with the inferred Dopaminergic tone given the data from either the ON and OFF acquisitions); 3) correlate the differences across frequency bins. To validate the correlation, we then randomize the power spectra by shuffling the frequency bins, to estimate a null distribution of the correlations given by chance, and use this null distribution to estimate the goodness of fit of our data.

## Data Availability

All data produced in the present study are available upon reasonable request to the authors

## Author contributions

Conceptualization and Methodology: MA, DD, MH, PS

Simulations and Data analysis: MA, DD, HA, MH, PS

Formal analysis: MA, DD, MH, PS

Coding: MA, MW

Resources: JR, RC, AE, VJ

Writing - Original Draft: MA, PS

Writing - Review - Editing: MA, DD, HA, JR, RC, MM, LC, PT, AZ, AE, VJ, PS

Visualization: MA, DD, HA, JR, RC, MM, LC, PT, AZ, AE, VJ, PS

Supervision and Project administration: DD, VJ, PS

Funding acquisition: VJ, PS All authors read and approved the final manuscript.

## Data availability statement

The datasets analysed in this study and the pipeline of the code are available from the corresponding author upon reasonable request.

## Supplementary figures

**Figure 6:**
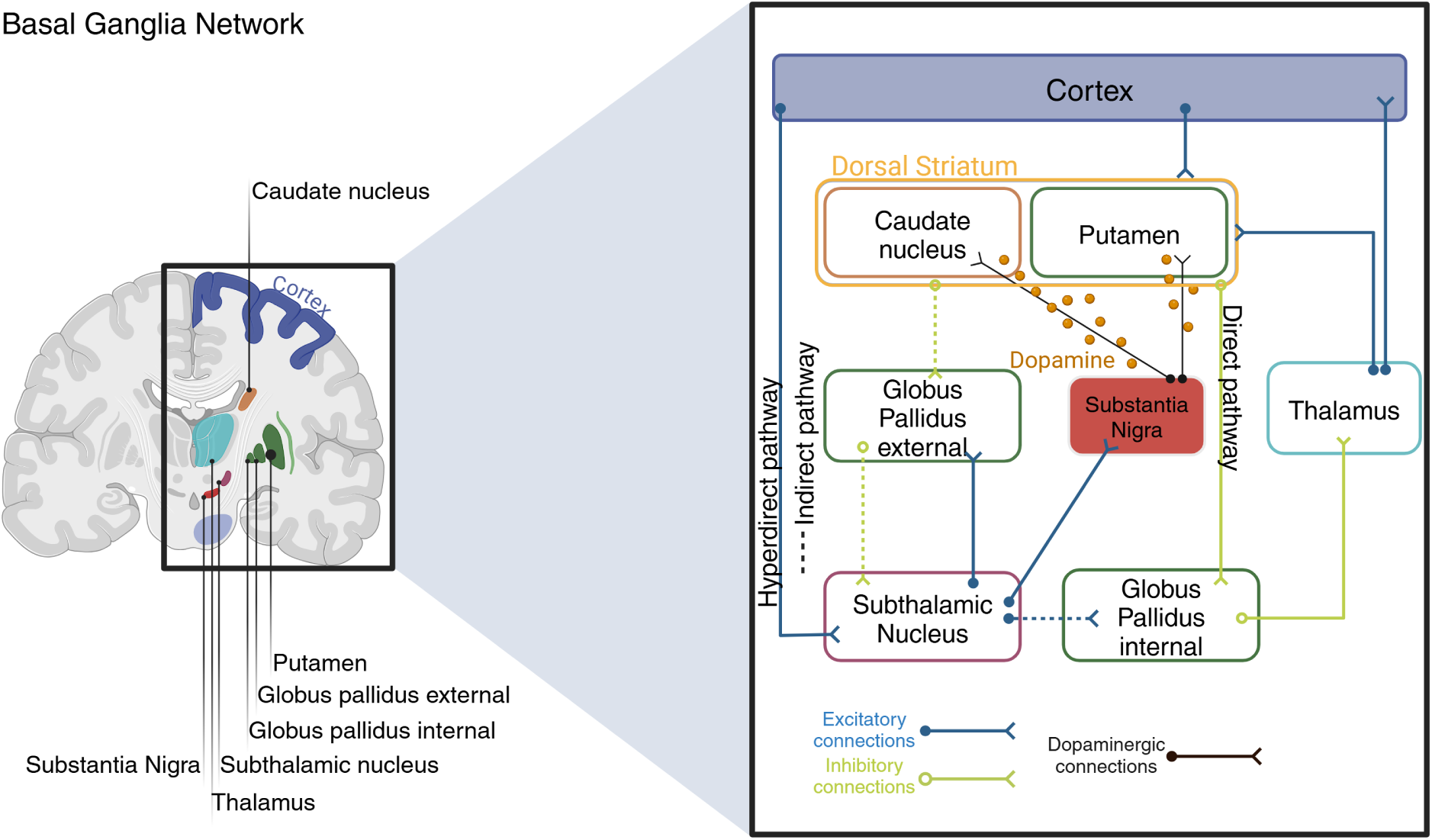
S1 Basal-ganglia neural networks. The picture shows a schematic diagram of the neural pathways in the basal ganglia. In this scheme, there are three main paths: the Hyperdirect Pathway connecting the Cortex to the STN, the Indirect Pathway and the Direct Pathway: Excitatory connec-tions are indicated by blue arrows while green arrows signify inhibitory connections. The dopaminergic connections are designed in black. Pictures created with BioRender.com

**Figure 7:**
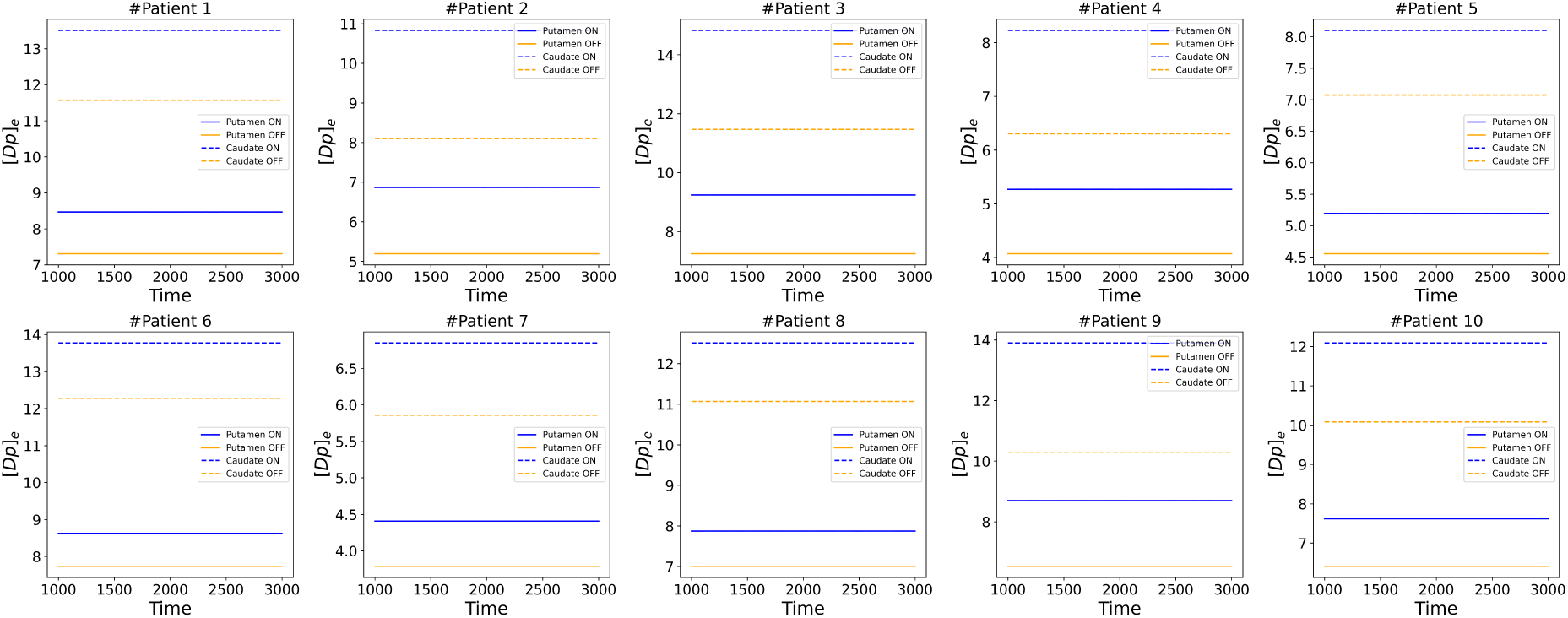
S2. Time series of the dopamine concentration in the Putamen and Caudate during the ON (blue) and OFF (orange) state.

## References

[1] A. Ascherio and M. A. Schwarzschild, “The epidemiology of parkinson’s disease: risk factors and prevention,” The Lancet Neurology, vol. 15, no. 12, pp. 1257–1272, 2016.

[2] W. Dauer and S. Przedborski, “Parkinson’s disease: mechanisms and models,” Neuron, vol. 39, no. 6, pp. 889–909, 2003.

[3] A. Horn, M. Reich, J. Vorwerk, N. Li, G. Wenzel, Q. Fang, T. Schmitz-Hübsch, R. Nickl, A. Kupsch, J. Volkmann, et al., “Connectivity predicts deep brain stimulation outcome in parkinson disease,” Annals of Neurology, vol. 82, no. 1, pp. 67–78, 2017.

[4] T. Merk, V. Peterson, W. J. Lipski, B. Blankertz, R. S. Turner, N. Li, A. Horn, R. M. Richardson, and W.-J. Neumann, “Electrocorticography is superior to subthalamic local field potentials for movement decoding in parkinson’s disease,” eLife, vol. 11, p. e75126, 2022.

[5] K. R. Chaudhuri and A. H. Schapira, “Non-motor symptoms of parkinson’s disease: dopaminergic pathophysiology and treatment,” The Lancet Neurology, vol. 8, no. 5, pp. 464–474, 2009.

[6] A. Tessitore, M. Cirillo, and R. De Micco, “Functional connectivity signatures of parkinson’s disease,” Journal of Parkinson’s Disease, vol. 9, pp. 637–652.

[7] M. Filippi, E. Sarasso, and F. Agosta, “Resting-state functional mri in parkinsonian syndromes,” Movement Disorders Clinical Practice, vol. 6, no. 2, pp. 104–117, 2019.

[8] P. Sorrentino, R. Rucco, F. Baselice, R. De Micco, A. Tessitore, A. Hillebrand, L. Mandolesi, M. Breakspear, L. L. Gollo, and G. Sorrentino, “Flexible brain dynamics underpins complex behaviours as observed in parkinson’s disease,” Scientific Reports, vol. 11, no. 1, p. 4051, 2021.

[9] H. Agouram, M. Neri, M. Angiolelli, D. Depannemaecker, J. Bahuguna, A. Schwey, J. Ŕegis, R. Car-ron, A. Eusebio, N. Malfait, et al., “L-dopa induced changes in aperiodic bursts dynamics relate to individual clinical improvement in parkinson’s disease,” medRxiv, 2024.

[10] D. Depannemaecker, C. Duprat, M. Angiolelli, C. S. Carbonell, H. Wang, S. Petkoski, P. Sorrentino, H. Sheheitli, and V. Jirsa, “A neural mass model with neuromodulation,” bioRxiv, 2024.

[11] P. Sorrentino, C. Seguin, R. Rucco, M. Liparoti, E. Troisi Lopez, S. Bonavita, M. Quarantelli, G. Sorrentino, V. Jirsa, and A. Zalesky, “The structural connectome constrains fast brain dynamics,” eLife, vol. 10, p. e67400, 2021.

[12] G. M. Duma, A. Danieli, G. Mento, V. Vitale, R. S. Opipari, V. Jirsa, P. Bonanni, and P. Sorrentino, “Altered spreading of neuronal avalanches in temporal lobe epilepsy relates to cognitive performance: A resting-state hdeeg study,” Epilepsia, vol. 64, no. 6, pp. 1278–1288, 2023.

[13] A. Romano, E. Troisi Lopez, L. Cipriano, M. Liparoti, R. Minino, A. Polverino, C. Cavaliere, M. Aiello, C. Granata, G. Sorrentino, et al., “Topological changes of fast large-scale brain dynamics in mild cognitive impairment predict early memory impairment: a resting-state, source reconstructed, magnetoencephalography study,” Neurobiology of Aging, vol. 132, pp. 36–46, 2023.

[14] M.-C. Corsi, P. Sorrentino, D. Schwartz, N. George, L. L. Gollo, S. Chevallier, L. Hugueville, A. E. Kahn, S. Dupont, D. S. Bassett, et al., “Measuring neuronal avalanches to inform brain-computer interfaces,” iScience, vol. 27, no. 2, p. 108734, 2024.

[15] K. A. Johnson and R. S. Goody, “The original michaelis constant: Translation of the 1913 michaelis–menten paper,” Biochemistry, vol. 50, no. 39, pp. 8264–8269, 2011.

[16] M. L. Kringelbach, J. Cruzat, J. Cabral, G. M. Knudsen, R. Carhart-Harris, P. C. Whybrow, N. K. Logothetis, and G. Deco, “Dynamic coupling of whole-brain neuronal and neurotransmitter systems,” Proceedings of the National Academy of Sciences, vol. 117, no. 17, pp. 9566–9576, 2020.

[17] E. Bullmore and O. Sporns, “Complex brain networks: graph theoretical analysis of structural and functional systems,” Nature Reviews Neuroscience, vol. 10, no. 3, pp. 186–198, 2009.

[18] K. Cranmer, J. Brehmer, and G. Louppe, “The frontier of simulation-based inference,” Proceedings of the National Academy of Sciences, vol. 117, no. 48, pp. 30055–30062, 2020.

[19] M. Hashemi, A. N. Vattikonda, V. Sip, M. Guye, F. Bartolomei, M. M. Woodman, and V. K. Jirsa, “The bayesian virtual epileptic patient: A probabilistic framework designed to infer the spatial map of epileptogenicity in a personalized large-scale brain model of epilepsy spread,” NeuroImage, vol. 217, p. 116839, 2020.

[20] A. Tejero-Cantero, J. Boelts, M. Deistler, J.-M. Lueckmann, C. Durkan, P. J. Goņcalves, D. S. Greenberg, and J. H. Macke, “sbi: A toolkit for simulation-based inference,” Journal of Open Source Software, vol. 5, no. 52, p. 2505, 2020.

[21] M. Hashemi, A. N. Vattikonda, J. Jha, V. Sip, M. M. Woodman, F. Bartolomei, and V. K. Jirsa, “Amortized bayesian inference on generative dynamical network models of epilepsy using deep neural density estimators,” Neural Networks, vol. 163, pp. 178–194, 2023.

[22] G. Papamakarios, T. Pavlakou, and I. Murray, “Masked autoregressive flow for density estimation,” in Advances in Neural Information Processing Systems, pp. 2338–2347, Curran Associates, Inc., 2017.

[23] P. J. Goņcalves, J.-M. Lueckmann, M. Deistler, M. Nonnenmacher, K. Ö cal, G. Bassetto, C. Chin-taluri, W. F. Podlaski, S. A. Haddad, T. P. Vogels, et al., “Training deep neural density estimators to identify mechanistic models of neural dynamics,” eLife, vol. 9, p. e56261, 2020.

[24] L. N. Ross and D. S. Bassett, “Causation in neuroscience: keeping mechanism meaningful,” Nature Reviews Neuroscience, vol. 25, no. 2, pp. 81–90, 2024.

[25] L. Chen and S. A. Campbell, “Exact mean-field models for spiking neural networks with adaptation,” arXiv preprint arXiv:2203.08341, 2022.

[26] U. Braun, A. Schaefer, R. F. Betzel, H. Tost, A. Meyer-Lindenberg, and D. S. Bassett, “From maps to multi-dimensional network mechanisms of mental disorders,” Neuron, vol. 97, no. 1, pp. 14–31, 2018.

[27] K. E. Stephan, S. Iglesias, J. Heinzle, and A. O. Diaconescu, “Translational perspectives for computational neuroimaging,” Neuron, vol. 87, no. 4, pp. 716–732, 2015.

[28] D. C. Van Essen and M. F. Glasser, “The human connectome project: Progress and prospects,” Cerebrum Dana Forum Brain Sci, vol. 2016, no. cer-10-16, 2016.

[29] H. Braak, E. Ghebremedhin, U. Rüb, H. Bratzke, and K. Del Tredici, “Stages in the development of parkinson’s disease-related pathology,” Cell and Tissue Research, vol. 318, no. 1, pp. 121–134, 2004.

[30] H. E. Wang, P. Triebkorn, M. Breyton, B. Dollomaja, J.-D. Lemarechal, S. Petkoski, P. Sorrentino, D. Depannemaecker, M. Hashemi, and V. K. Jirsa, “Virtual brain twins: from basic neuroscience to clinical use,” National Science Review, vol. 11, no. nwae079, 2024.

[31] P. Sorrentino, E. Troisi Lopez, A. Romano, C. Granata, M.-C. Corsi, G. Sorrentino, and V. Jirsa, “Brain fingerprint is based on the aperiodic, scale-free, neuronal activity,” NeuroImage, vol. 277, p. 120260, 2023.

[32] Y. Yu, D. Escobar Sanabria, J. Wang, C. M. Hendrix, J. Zhang, S. D. Nebeck, A. M. Amundson, Z. B. Busby, D. L. Bauer, M. D. Johnson, et al., “Parkinsonism alters beta burst dynamics across the basal ganglia–motor cortical network,” Journal of Neuroscience, vol. 41, no. 10, pp. 2274–2286, 2021.

[33] G. Tinkhauser, A. Pogosyan, H. Tan, D. M. Herz, A. A. Kühn, and P. Brown, “Beta burst dynamics in parkinson’s disease off and on dopaminergic medication,” Brain: A Journal of Neurology, vol. 140, no. 11, pp. 2968–2981, 2017.

[34] R. Lofredi, L. Okudzhava, F. Irmen, C. Brücke, J. Huebl, J. K. Krauss, G.-H. Schneider, K. Faust, W.-J. Neumann, and A. A. Kühn, “Subthalamic beta bursts correlate with dopamine-dependent motor symptoms in 106 parkinson’s patients,” NPJ Parkinson’s Disease, vol. 9, no. 1, pp. 1–9, 2023.

[35] K. A. M. Pauls, O. Korsun, J. Nenonen, J. Nurminen, M. Liljestrőm, J. Kujala, E. Pekkonen, and H. Renvall, “Cortical beta burst dynamics are altered in parkinson’s disease but normalized by deep brain stimulation,” NeuroImage, vol. 257, p. 119308, 2022.

[36] M. C. Vinding, P. Tsitsi, J. Waldthaler, R. Oostenveld, M. Ingvar, P. Svenningsson, and D. Lundqvist, “Reduction of spontaneous cortical beta bursts in parkinson’s disease is linked to symptom severity,” Brain Communications, vol. 2, no. 2, p. fcaa052, 2020.

[37] S. Scarpetta, N. Morisi, C. Mutti, N. Azzi, I. Trippi, R. Ciliento, I. Apicella, G. Messuti, M. Angi-olelli, F. Lombardi, et al., “Criticality of neuronal avalanches in human sleep and their relationship with sleep macro-and micro-architecture,” iScience, vol. 26, no. 6, p. 107840, 2023.

[38] M. Betancourt, “Calibrating model-based inferences and decisions,” arXiv preprint arXiv:1805.08354, 2018.

[39] A. Pavlides, S. J. Hogan, and R. Bogacz, “Computational models describing possible mechanisms for generation of excessive beta oscillations in parkinson’s disease,” PLoS Computational Biology, vol. 11, no. 12, p. e1004609, 2015.

[40] A. Oswal, C. Cao, C.-C. Yeh, W.-J. Neumann, J. Gratwicke, H. Akram, A. Horn, D. Li, S. Zhan, C. Zhang, et al., “Neural signatures of hyperdirect pathway activity in parkinson’s disease,” Nature Communications, vol. 12, no. 1, pp. 1–15, 2021.

[41] G. Deco, A. Ponce-Alvarez, D. Mantini, G. L. Romani, P. Hagmann, and M. Corbetta, “Resting-state functional connectivity emerges from structurally and dynamically shaped slow linear fluctuations,” Journal of Neuroscience, vol. 33, no. 27, pp. 11239–11252, 2013.

[42] X. Kong, R. Kong, C. Orban, P. Wang, S. Zhang, K. Anderson, A. Holmes, J. D. Murray, G. Deco, M. van den Heuvel, et al., “Sensory-motor cortices shape functional connectivity dynamics in the human brain,” Nature Communications, vol. 12, no. 1, p. 6373, 2021.

[43] J. M. Meier, D. Perdikis, A. Blickensdőrfer, L. Stefanovski, Q. Liu, O. Maith, H. Ü . Dinkelbach, J. Baladron, F. H. Hamker, and P. Ritter, “Virtual deep brain stimulation: Multiscale co-simulation of a spiking basal ganglia model and a whole-brain mean-field model with the virtual brain,” Experimental Neurology, vol. 354, p. 114111, 2022.

[44] V. M. Saenger, J. Kahan, T. Foltynie, K. Friston, T. Z. Aziz, A. L. Green, T. J. van Hartevelt, J. Cabral, A. B. A. Stevner, H. M. Fernandes, et al., “Uncovering the underlying mechanisms and whole-brain dynamics of deep brain stimulation for parkinson’s disease,” Scientific Reports, vol. 7, no. 1, p. 9882, 2017.

[45] M. D. Humphries, J. A. Obeso, and J. K. Dreyer, “Insights into parkinson’s disease from computational models of the basal ganglia,” *Journal of Neurology*, Neurosurgery & Psychiatry, vol. 89, no. 11, pp. 1181–1188, 2018.

[46] R. Oostenveld, P. Fries, E. Maris, and J.-M. Schoffelen, “Fieldtrip: Open source software for advanced analysis of meg, eeg, and invasive electrophysiological data,” Computational Intelligence and Neuroscience, vol. 2011, p. 1, 2011.

[47] G. Niso, L. R. Krol, E. Combrisson, A.-S. Dubarry, M. A. Elliott, C. Fraņcois, Y. Héjja-Brichard, S. K. Herbst, K. Jerbi, V. Kovic, et al., “Good scientific practice in eeg and meg research: Progress and perspectives,” NeuroImage, vol. 257, p. 119056, 2022.

[48] D. C. Van Essen, S. M. Smith, D. M. Barch, T. E. Behrens, E. Yacoub, K. Ugurbil, and W.-M. H. Consortium, “The wu-minn human connectome project: an overview,” NeuroImage, vol. 80, pp. 62–79, 2013.

[49] M. F. Glasser, S. N. Sotiropoulos, J. A. Wilson, T. S. Coalson, B. Fischl, J. L. Andersson, J. Xu, S. Jbabdi, M. Webster, J. R. Polimeni, et al., “The minimal preprocessing pipelines for the human connectome project,” NeuroImage, vol. 80, pp. 105–124, 2013.

[50] J. Tournier, R. Smith, D. Raffelt, R. Tabbara, T. Dhollander, M. Pietsch, D. Christiaens, B. Jeurissen, C.-H. Yeh, and A. Connelly, “Mrtrix3: A fast, flexible and open software framework for medical image processing and visualisation,” NeuroImage, vol. 202, p. 116137, 2019.

[51] B. Jeurissen, J. Tournier, T. Dhollander, A. Connelly, and J. Sijbers, “Multi-tissue constrained spherical deconvolution for improved analysis of multi-shell diffusion mri data,” NeuroImage, vol. 103, pp. 411–426, 2014.

[52] D. Raffelt, T. Dhollander, J. Tournier, R. Tabbara, R. Smith, E. Pierre, and A. Connelly, “Bias field correction and intensity normalisation for quantitative analysis of apparent fibre density,” Preprint, 2017.

[53] R. E. Smith, J. Tournier, F. Calamante, and A. Connelly, “Anatomically-constrained tractography: Improved diffusion mri streamlines tractography through effective use of anatomical information,” NeuroImage, vol. 62, no. 3, pp. 1924–1938, 2012.

[54] R. E. Smith, J. Tournier, F. Calamante, and A. Connelly, “Sift2: Enabling dense quantitative assessment of brain white matter connectivity using streamlines tractography,” NeuroImage, vol. 119, pp. 338–351, 2015.

[55] A. Horn, N. Li, T. A. Dembek, A. Kappel, C. Boulay, S. Ewert, A. Tietze, A. Husch, T. Perera, W.-J. Neumann, et al., “Lead-dbs v2: Towards a comprehensive pipeline for deep brain stimulation imaging,” NeuroImage, vol. 184, pp. 293–316, 2019.

[56] S. Ewert, P. Plettig, N. Li, M. M. Chakravarty, D. L. Collins, T. M. Herrington, A. A. Kühn, and A. Horn, “Toward defining deep brain stimulation targets in mni space: A subcortical atlas based on multimodal mri, histology and structural connectivity,” NeuroImage, vol. 170, pp. 271–282, 2018.

[57] R. S. Desikan, F. Śegonne, B. Fischl, B. T. Quinn, B. C. Dickerson, D. Blacker, R. L. Buckner, A. M. Dale, R. P. Maguire, B. T. Hyman, et al., “An automated labeling system for subdividing the human cerebral cortex on mri scans into gyral based regions of interest,” NeuroImage, vol. 31, no. 3, pp. 968–980, 2006.

[58] B. Fischl, “Freesurfer,” NeuroImage, vol. 62, no. 2, pp. 774–781, 2012.

[59] A. Gramfort, M. Luessi, E. Larson, D. A. Engemann, D. Strohmeier, C. Brodbeck, R. Goj, M. Jas, T. Brooks, L. Parkkonen, et al., “Meg and eeg data analysis with mne-python,” Frontiers in Neuroscience, vol. 7, p. 267, 2013.

[60] M. R. DeLong, “Primate models of movement disorders of basal ganglia origin,” Trends in Neurosciences, vol. 13, no. 7, pp. 281–285, 1990.

[61] A. Roberts, “Modify the improved euler scheme to integrate stochastic differential equations,” 2012.

[62] A. Hussain, “Numerical solutions of stochastic differential equations by using heun’s method,” 2023.

[63] A. Gelman, J. B. Carlin, H. S. Stern, and D. B. Rubin, Bayesian Data Analysis. Chapman and Hall/CRC, 1995.

[64] “Bayesian statistics and modelling,” Nature Reviews Methods Primers, 2020.

[65] I. Kobyzev, S. J. Prince, and M. A. Brubaker, “Normalizing flows: An introduction and review of current methods,” IEEE Transactions on Pattern Analysis and Machine Intelligence, vol. 43, no. 11, pp. 3964–3979, 2021.

[66] A. Gelman, A. Vehtari, D. Simpson, C. C. Margossian, B. Carpenter, Y. Yao, L. Kennedy, J. Gabry, P.-C. Bürkner, and M. Modŕak, “Bayesian workflow,” arXiv preprint arXiv:, 2020.

